# Parental smoking and respiratory outcomes in young childhood cancer survivors

**DOI:** 10.1101/2024.05.31.24308191

**Authors:** Maša Žarković, Grit Sommer, Carina Nigg, Tomáš Sláma, Christine Schneider, Marc Ansari, Nicolas von der Weid, Christina Schindera, Claudia E Kuehni

**Author notes:** **Corresponding author** Claudia Kuehni, Childhood Cancer Research Group, Institute of Social and Preventive Medicine, University of Bern, Mittelstrasse 43, 3012 Bern, Switzerland.

## Abstract

**Background:** Passive exposure to cigarette smoke has negative effects on respiratory health. Childhood cancer survivors (CCS) are at an increased risk for respiratory disease due to treatment regimens that may harm the respiratory system. The objective of this study was to assess the prevalence of parental smoking among CCS and investigate its association with respiratory outcomes.

**Procedure:** As part of the Swiss Childhood Cancer Survivor Study, between 2007 and 2022 we sent questionnaires to parents of children aged ≤16 years who had survived ≥ 5 years after cancer diagnosis. Parents reported on their children’s respiratory outcomes including recurrent upper respiratory tract infections (otitis media and sinusitis), asthma, and lower respiratory symptoms (chronic cough persisting > 3 months, current and exercise wheeze), and on parental smoking. We used multivariable logistic regression to investigate associations between parental smoking and respiratory outcomes.

**Results:** Our study included 1037 CCS (response rate 66%). Median age at study was 12 years (interquartile range [IQR] 10–14). Eighteen percent of mothers and 23% of fathers reported current smoking. CCS exposed to smoking mothers were more likely to have recurrent upper respiratory tract infections (OR 2.1; 95% CI 1.1–3.7) and lower respiratory symptoms (OR 2.0; 95%CI 1.1-3.7). We found no association with paternal smoking.

**Conclusions:** A substantial proportion of CCS in Switzerland have parents who smoke. Exposure to maternal smoking was associated with higher prevalence of upper and lower respiratory problems. Physicians should advise and assist families of CCS in their endeavors to quit smoking.

## Introduction

Childhood cancer survivors (CCS) are at increased risk of pulmonary late effects because of their cancer treatment [1]. Chemotherapy, radiotherapy and surgery involving the chest, and hematopoietic stem cell transplantation may affect lung function and cause symptoms such as chronic cough and exercise-induced shortness of breath [2, 3]. Cumulative incidence of adverse pulmonary outcomes continues to increase years after completing cancer treatment [4]. This indicates continuing susceptibility of survivors’ lungs to initiation and exacerbation of pulmonary diseases. Cancer therapies may also result in delayed immune reconstitution, which leaves patients vulnerable to infections well beyond the conclusion of their treatment [5, 6]. Consequently, infectious complications, such as upper respiratory tract infections including sinusitis and otitis media, remain an important cause of late morbidity among survivors, impacting their health and increasing hospitalization rates [6, 7].

Even among otherwise healthy individuals, environmental tobacco smoke is a risk factor for respiratory symptoms and infections. Children exposed to parental tobacco smoke have increased susceptibility to recurrent otitis media, asthma, cough, wheeze, and lower respiratory illnesses [8]. Globally, nearly 40% of children are exposed to tobacco smoke [9]. In Switzerland, almost one-third of children (31%) grow up in homes in which a parent smokes [10]. Parental smoking may have persistent effects on the respiratory systems of children with potential consequences continuing into adulthood [11]. While this is known for the general population, research on the particularly vulnerable group of CCS is lacking. To address this, we examined the prevalence of parental smoking among Swiss CCS aged ≤16 years in a nationwide study and investigated associations between parental smoking and respiratory outcomes.

## Methods

### The Swiss Childhood Cancer Survivor Study (SCCSS)

The SCCSS is a population-based, long-term follow-up study (www.swiss-ccss.ch) of survivors registered in the Swiss Childhood Cancer Registry (ChCR, www.kinderkrebsregister.ch) [12]. The ChCR is a national registry founded in 1976 that includes all Swiss residents diagnosed before the age of 20 years with leukemias, lymphomas, central nervous system (CNS) tumors, malignant solid tumors, or Langerhans cell histiocytosis [13]. As part of the SCCSS, we sent questionnaires to former cancer patients who survived for five years or more since their initial diagnosis. Detailed methods of the SCCSS have been published elsewhere [12]. For this study, we analyzed questionnaires returned between 2007 and 2022 by parents of CCS younger than 16 years who had answered questions on smoking status and their child’s respiratory problems. The Ethics Committee of the Canton of Bern granted ethical approval to the ChCR and the SCCSS (166/2014; 2021-0146).

### Definition of respiratory outcomes

The questionnaires assessed smoking within the family, and children’s respiratory symptoms and infections occurring during the previous 12 months. Questions inquired about the presence of asthma (yes/no), chronic cough lasting longer than three months (yes/no), recurrent sinusitis (yes/no), and recurrent otitis media (yes/no) in CCS. The questionnaires sent after 2015 also asked whether CCS ever had wheezed in their life (yes/no), in the last 12 months (yes/no), and during exercise (yes/no). From those questions, we created three main groups of respiratory outcomes for the analyses: recurrent upper respiratory tract infections (URTI), asthma, and lower respiratory symptoms. Participants with recurrent URTI included those who reported either recurrent sinusitis or recurrent otitis media, or both. Participants with lower respiratory symptoms included those who had either chronic cough, at least one episode of wheeze in the past 12 months, or wheeze during exercise. The original questions are available upon request.

### Explanatory variables

### Parental smoking

We asked mothers and fathers separately if they had ever smoked. Possible answers were “no, never”, “yes, but stopped”, and “yes, still smoke today”. If data was missing for both the maternal and paternal smoking status, participants were excluded from further analyses. We coded parental smoking as a binary variable (yes/no) where “yes” indicated current smoker and “no” indicated former or never smoker.

### Sociodemographic characteristics

We assessed the following sociodemographic characteristics of CCS: age at study, sex, Swiss language region (German, French, or Italian), living situation, parental nationality, education, and employment status. We classified the living situation of CCS as living with both parents, only with the mother, only with the father, or in other circumstances (in a children’s home or with relatives). Parents’ nationality was categorized as Swiss or other. We considered survivors as having a migration background if they had at least one parent whose nationality was other than Swiss. We divided the highest level of education parents attained into three categories: primary schooling (compulsory schooling only, ≤ 9 years), secondary education (vocational training or upper secondary education), or tertiary education (university or technical college education).

### Clinical characteristics

We received the following cancer-related variables from the ChCR: age at diagnosis, time since diagnosis, chemotherapy (yes/no), radiotherapy (yes/no), surgery (yes/no), hematopoietic stem cell transplantation (yes/no), relapse status (yes/no), and diagnosis coded according to the International Classification of Childhood Cancer, third edition (ICCC-3) [14].

### Statistical analysis

We used medians and interquartile ranges (IQR) for continuous variables and numbers and percentages for categorical variables to describe sociodemographic and clinical characteristics of survivors and parental smoking status. To examine the association between parental smoking and respiratory outcomes, we used logistic regression. First, we fit univariable logistic regression models to identify sociodemographic and clinical factors associated with recurrent URTI, asthma, and lower respiratory symptoms. Exposure variables that showed an association with the outcome variables at a level of p<0.1 were included in a multivariable logistic regression model. We incorporated maternal and paternal smoking, sex, and age a priori. All respiratory outcomes had a low number of missing answers (<10%), so we assumed that the respective respiratory disease or symptom was absent. If there were missing values in other independent or dependent variables, complete case analysis was performed. We used Stata (Version 16.1, Stata Corporation, Austin, TX) for all analyses.

## Results

### Characteristics of the study population

Of the 1846 eligible survivors identified through the Swiss Childhood Cancer Registry, we sent the parents of 1628 a questionnaire. Questionnaires were returned by parents in 1070 families (a response rate of 66%). We excluded questionnaires of 33 survivors for whom we found no answers to any of the questions on parental smoking or on respiratory outcomes. The final dataset used for the analysis thus included 1037 children (Figure 1). Their median age at study was 12 years (interquartile range [IQR] 10-14), median age at diagnosis was 3 years (IQR 2-5), and median time since diagnosis 8 years (IQR 7-10). The most common diagnoses were leukemia (39%), CNS tumors (16%), and neuroblastoma (9%) (Table 1).

**FIGURE 1.**
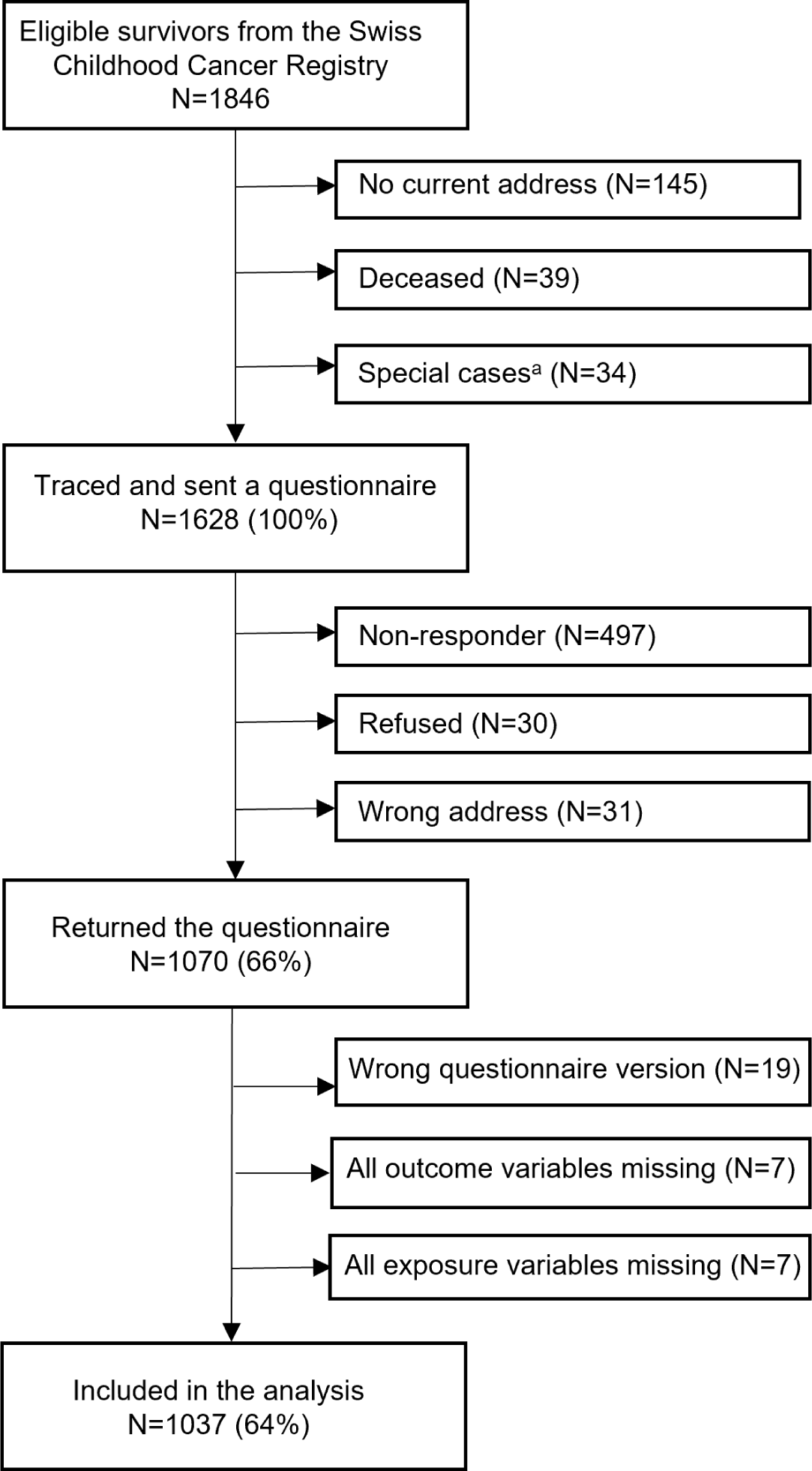
Flow diagram of the study population starting from eligible survivors identified by the Swiss Childhood Cancer Registry to those included in the analysis. ^a^ Special cases were identified after contacting clinics where survivors had been treated to evaluate eligibility and to exclude patients with current conditions that preclude contact (e.g., recent relapse, palliative situation)

**TABLE 1.**
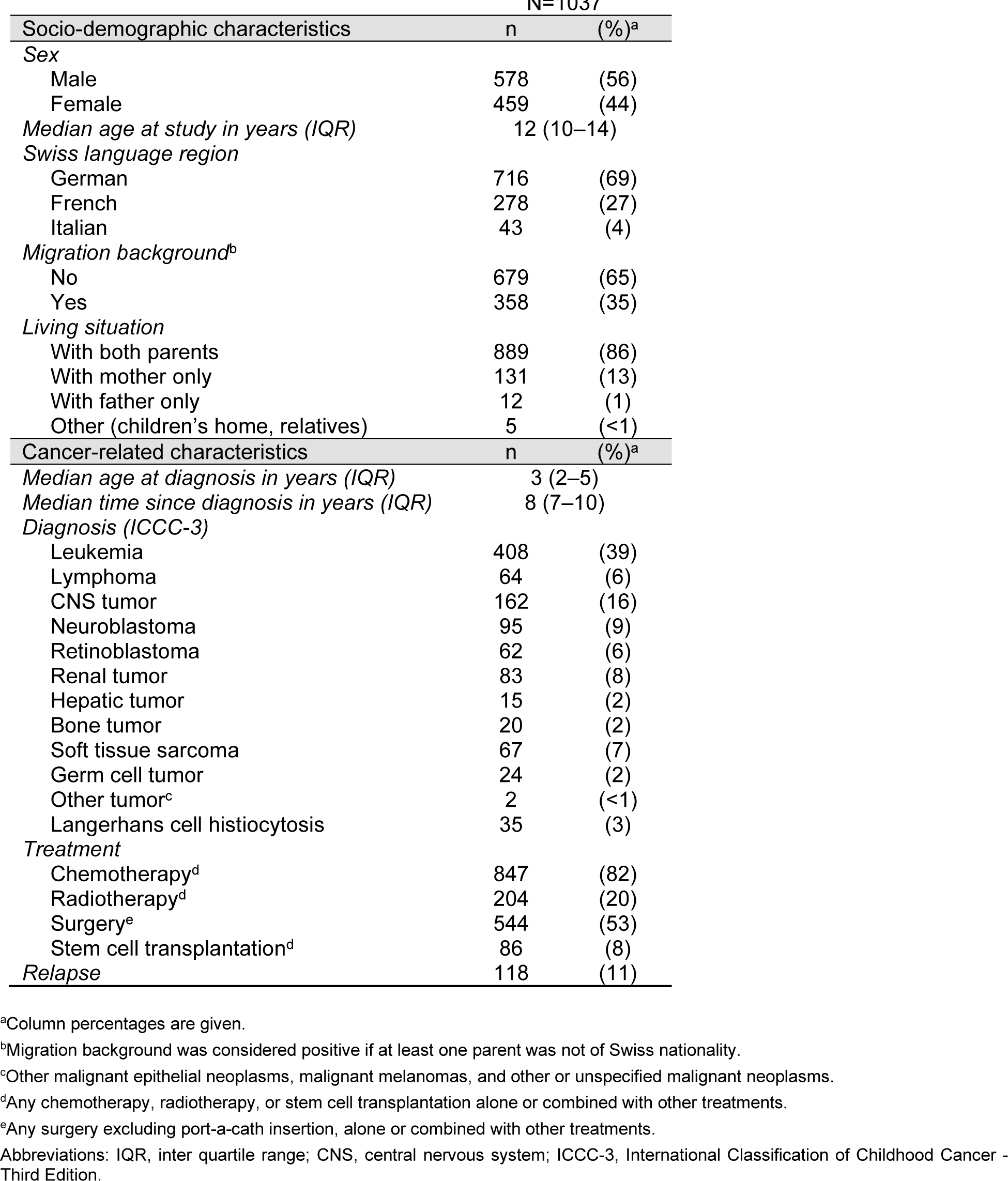
Characteristics of the study population.

| N=1037 |  |  |
| --- | --- | --- |
| Socio-demographic characteristics | n | (%)<sup>a</sup> |
| **Sex** |  |  |
| Male | 578 | (56) |
| Female | 459 | (44) |
| *Median age at study in years (IQR)* | 12 (10–14) |  |
| *Swiss language region* |  |  |
| German | 716 | (69) |
| French | 278 | (27) |
| Italian | 43 | (4) |
| *Migration background<sup>b</sup>* |  |  |
| No | 679 | (65) |
| Yes | 358 | (35) |
| *Living situation* |  |  |
| With both parents | 889 | (86) |
| With mother only | 131 | (13) |
| With father only | 12 | (1) |
| Other (children's home, relatives) | 5 | (<1) |
| Cancer-related characteristics | n | (%)<sup>a</sup> |
| *Median age at diagnosis in years (IQR)* | 3 (2–5) |  |
| *Median time since diagnosis in years (IQR)* | 8 (7–10) |  |
| *Diagnosis (ICCC-3)* |  |  |
| Leukemia | 408 | (39) |
| Lymphoma | 64 | (6) |
| CNS tumor | 162 | (16) |
| Neuroblastoma | 95 | (9) |
| Retinoblastoma | 62 | (6) |
| Renal tumor | 83 | (8) |
| Hepatic tumor | 15 | (2) |
| Bone tumor | 20 | (2) |
| Soft tissue sarcoma | 67 | (7) |
| Germ cell tumor | 24 | (2) |
| Other tumor<sup>c</sup> | 2 | (<1) |
| Langerhans cell histiocytosis | 35 | (3) |
| *Treatment* |  |  |
| Chemotherapy<sup>d</sup> | 847 | (82) |
| Radiotherapy<sup>d</sup> | 204 | (20) |
| Surgery<sup>e</sup> | 544 | (53) |
| Stem cell transplantation<sup>d</sup> | 86 | (8) |
| *Relapse* | 118 | (11) |

<sup>a</sup>Column percentages are given.
<sup>b</sup>Migration background was considered positive if at least one parent was not of Swiss nationality.
<sup>c</sup>Other malignant epithelial neoplasms, malignant melanomas, and other or unspecified malignant neoplasms.
<sup>d</sup>Any chemotherapy, radiotherapy, or stem cell transplantation alone or combined with other treatments.
<sup>e</sup>Any surgery excluding port-a-cath insertion, alone or combined with other treatments.
Abbreviations: IQR, inter quartile range; CNS, central nervous system; ICC-3, International Classification of Childhood Cancer - Third Edition.

Among parents, median age at study was 43 years for mothers (IQR 40-47) and 46 years for fathers (IQR 42-50) (Supplemental Table S1). Secondary education was the most frequently attained education level of mothers (50%), while tertiary education was the level achieved by the highest proportion of fathers (49%). Almost all survivors lived with both parents (86%) (Table 1). Most fathers (80%), but only 10% of mothers worked full-time (Supplemental Table S1).

### Prevalence of parental smoking

Among mothers, 18% indicated that they currently smoked (95% confidence interval [CI] 15-20%), 22% said they were former smokers who had stopped before the study (95%CI 19-25%), and 60% had never smoked (95%CI 57-63%) (Supplemental Table S1). Among fathers, 24% were active smokers at the time of the study (95%CI 21-27%), 24% were former smokers (95%CI 22-27%), and 52% had never smoked (95%CI 49-55%). In 11% of families, both parents smoked (95%CI 9-13%), and in 20% only one parent was an active smoker (95%CI 18-23%). In most families, none of the parents smoked (69%, 95%CI 66-72%) (Supplemental Figure S1). Smoking was more common in parents with a lower educational level, and among unemployed fathers (Supplemental Table S2).

### Prevalence of respiratory outcomes

Eight percent of survivors reported recurrent upper respiratory tract infections (95%CI 7-10%), while asthma was reported by 5% (95%CI 4-7%) (Table 2). In total, 6% of survivors reported lower respiratory symptoms (95%CI 5-8%), including 2% with chronic cough (95%CI 1-3%), 6% with wheeze (95%CI 4-8%), and 6% with wheeze during exercise (95%CI 4-8%). Almost one-third of the survivors with lower respiratory symptoms (32%) reported more than one of these symptoms.

**TABLE 2.** Prevalence of upper and lower respiratory outcomes in childhood cancer survivors.

| N=1037 |  |  |  |
| --- | --- | --- | --- |
|  | n | % | (95%CI) |
| **Recurrent URTI<sup>a</sup>** | 85 | 8 | (7–10) |
| Recurrent otitis<sup>b</sup> | 54 | 5 | (4–7) |
| Recurrent sinusitis | 35 | 3 | (2–5) |
| **Asthma** | 54 | 5 | (4–7) |
| **Lower respiratory symptoms<sup>c</sup>** | 65 | 6 | (5–8) |
| Chronic cough | 22 | 2 | (1–3) |
| Current wheeze<sup>d</sup> | 33 | 6 | (4–8) |
| Wheeze during exercise<sup>d</sup> | 32 | 6 | (4–8) |

<sup>a</sup>Defined as having reported at least one of the following recurrent infections
<sup>b</sup>Question not asked in a part of questionnaires (total N=1003)
<sup>c</sup>Defined as having reported at least one of the following symptoms
<sup>d</sup>Question not asked in a part of questionnaires. The percentages are derived from the participants that were asked this question (total N=569)
Abbreviation: URTI, upper respiratory tract infections; CI, confidence interval

### Associations between parental smoking and respiratory outcomes

In multivariable regression, survivors whose mothers smoked were more likely to have recurrent URTI (odds ratio [OR] 2.1, 95%CI 1.1-3.7) and lower respiratory symptoms (OR 2.0, 95%CI 1.1-3.7) compared to survivors whose mothers did not smoke (Figure 2, Supplemental Table S3). Paternal smoking was unrelated to recurrent URTI, asthma, and lower respiratory symptoms.

**FIGURE 2.**
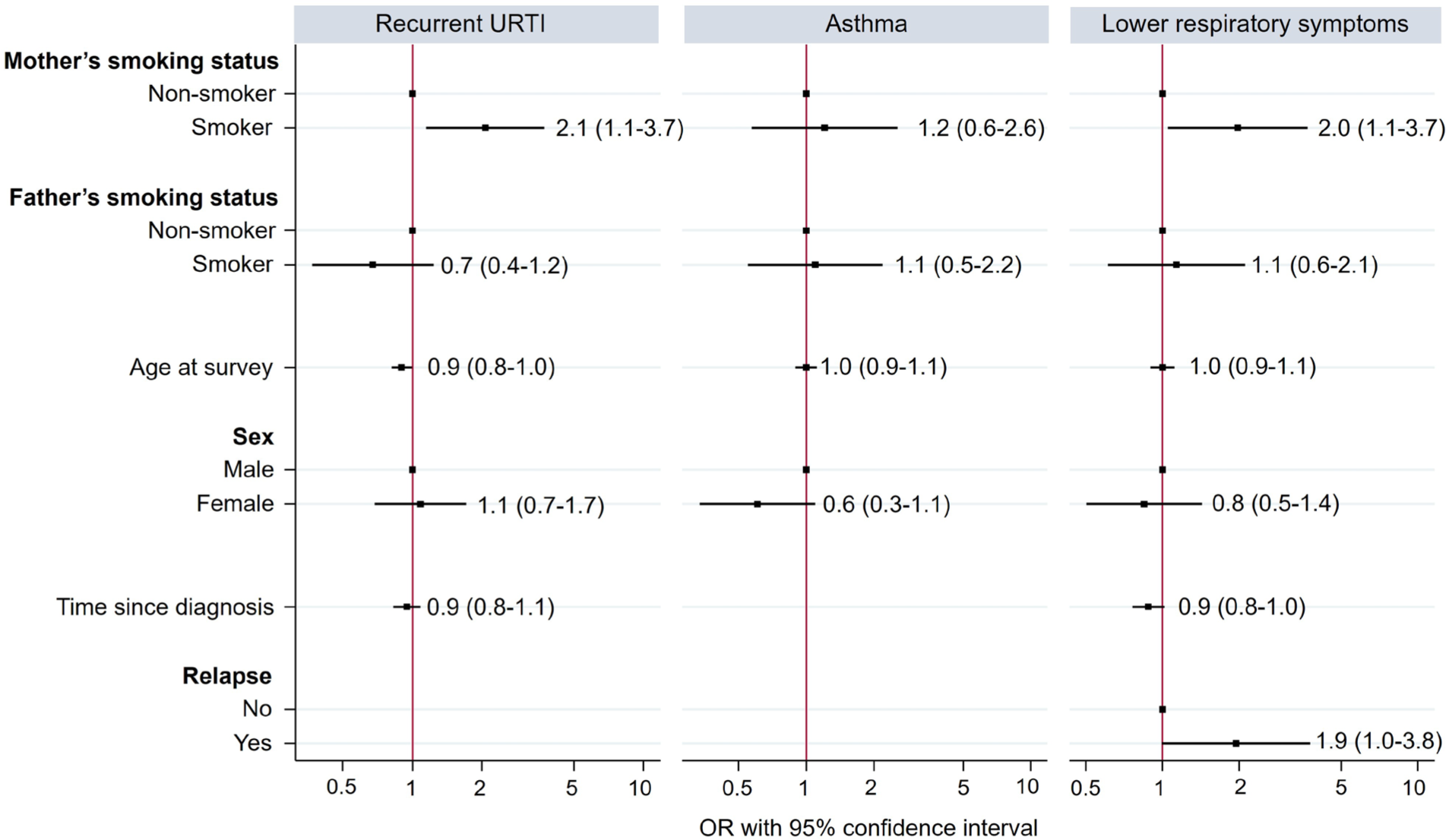
Associations between parental smoking and respiratory outcomes in childhood cancer survivors. Multivariable logistic regression models adjusted for age at survey, sex, time since diagnosis, and relapse. Abbreviation: URTI, upper respiratory tract infections; OR, odds ratio.

Older survivors were less likely have recurrent URTI (OR 0.9, 95%CI 0.8-1.0). With increasing time since diagnosis, survivors were less likely to have recurrent URTI in the univariable regression (OR 0.9, 95%CI 0.8-1.0), but this effect was not seen after adjusting in the multivariable regression (OR 0.9, 95%CI 0.8-1.1). Survivors with a history of relapse were more likely to have lower respiratory symptoms (OR 1.9, 95%CI 1.0-3.8). Sex was not associated with any of the respiratory outcomes. Detailed results from univariable logistic regression can be found in Supplemental Table S4.

## Discussion

A substantial proportion of young childhood cancer survivors in this nationwide, population-based cohort study is exposed to parents who smoke (31%). We found that maternal smoking in these families was associated with two-fold increased risk of recurrent upper respiratory tract infections and lower respiratory symptoms.

### Prevalence of parental smoking

Despite the general decline in smoking worldwide over the past two decades [15, 16], our study found that a significant proportion of Swiss CCS are exposed to parental smoking, with 18% of mothers and 24% of fathers reported as smokers. A school-based survey on children’s respiratory health conducted in the Canton of Zurich between 2013 and 2016 found slightly higher rates, with 20% of mothers and 30% of fathers smoking [17]. Similarly, a 2017 nationwide report on tobacco use by the Swiss Federal Statistical Office indicated that 23% of women and 31% of men aged 15 and older were daily or occasional smokers [16]. Yet little is known about parental smoking in the families of CCS. A study conducted in the USA found that only 6% of parents of CCS were active smokers compared to 12% of adults in the general population [18, 19]. The lower prevalence among parents of CCS may be because parents of survivors tend to adopt more health-conscious behaviors [18]. However, it could also be that underreporting is more common in this population.

### Associations with respiratory outcomes

Maternal smoking was associated with a greater risk of recurrent upper respiratory tract infections and lower respiratory symptoms among young (<16 years) childhood cancer survivors in our study. These results are in line with questionnaire-based studies in the general population that consistently have found a stronger impact of maternal smoking on respiratory health than paternal smoking [20–23]. Women are more often primary caregivers and spend more time on average at home with their children [24]. In our study, almost all of the CCS lived with both parents (86%) or only their mothers (13%), and there was a pronounced difference between employment status of parents, with only 10% of mothers having full-time jobs compared to 80% of fathers. This suggests that survivors may be more susceptible to the adverse effects of maternal smoking due to increased exposure to tobacco smoke within the household environment.

Passive smoking contributes to respiratory morbidity in children [8]. In the late 1990s, a series of comprehensive systematic reviews with meta-analyses revealed consistent evidence for associations between maternal smoking and recurrent otitis media (OR 1.48, 95%CI 1.08-2.04) [25], wheeze (OR 1.28, 95% CI 1.19-1.38) and cough (OR 1.40, 95%CI 1.20-1.64). [26] A multicountry, questionnaire-based study of 220 407 children (6-7 years) and 350 654 adolescents (13-14 years) found that maternal smoking was associated with wheeze both in children (OR 1.28, 95%CI 1.22-1.34) and adolescents (OR 1.32, 95%CI 1.26-1.37) [21]. Compared to those results, our observations in CCS indicated slightly stronger associations between maternal smoking and recurrent upper respiratory tract infections and lower respiratory symptoms. Survivors with a history of relapse were more likely to experience lower respiratory symptoms, perhaps because of more intense and prolonged treatments including chemotherapy, radiation, and hematopoietic stem cell transplantation [27, 28].

In contrast to previous studies conducted among the general population [21–23, 29, 30], we found no association between parental smoking and asthma. A systematic review found that children whose parents smoked were more likely to have asthma than children with nonsmoking parents (OR 1.21, 95%CI 1.10-1.34) [31]. The lack of association observed in our study may be due to underreporting or underdiagnosing of asthma. Several studies have found that children with parents who smoke were less likely to receive an asthma diagnosis [32, 33]. This could be because parents who smoke are less likely to seek medical advice regarding mild respiratory symptoms that could be related to smoking in the family [32, 33]. A comprehensive literature summary of previous studies can be found in the supplement (Supplemental literature summary).

### Limitations and strengths

The cross-sectional design of this study does not allow causal conclusions regarding smoking and respiratory outcomes. However, the biological plausibility connecting passive smoking and respiratory conditions is well established [8]. Passive tobacco smoke has toxic effects on the mucosa, impairs ciliary function and local immune defenses, and results in prolonged inflammation and susceptibility to infections such as otitis media and sinusitis [34]. It also contributes to airway thickening and irritation thereby predisposing children to respiratory symptoms including wheeze and cough [35]. Self-reported exposures and outcomes may have been subject to recall bias and led to misclassification of some respiratory outcomes. Our study also had limited statistical power because of the low number of respiratory events. Therefore, we could not determine associations between parental smoking and respiratory outcomes within different diagnostic and cancer treatment subgroups.

The strengths of our study include the population-based design and a high response rate, which ensured that the study population is representative of CCS in Switzerland. We used high quality clinical information based on medical records from the Swiss Childhood Cancer Registry and assessed a diverse spectrum of respiratory outcomes.

In conclusion, our study found that exposure to parental smoking is associated with increased risk of respiratory symptoms and infections among Swiss childhood cancer survivors. Further study of smoking in families of CCS is needed and would benefit from a longitudinal design and a control group of children without a cancer history. In the meantime, particularly given definitive evidence of the harm of passive smoking, the integration of parental smoking cessation strategies in childhood cancer patients’ medical and psychosocial care is essential to create a healthier home environment and promote respiratory health in this vulnerable population.

Conflict of interest statement

The authors declare there is no conflict of interest related to the study.

## Data Availability

All data produced in the present study are available upon reasonable request to the authors

## List of abbreviations

CCS: Childhood Cancer Survivor
SCCSS: Swiss Childhood Cancer Survivor Study
ChCR: Swiss Childhood Cancer Registry
CNS: Central Nervous System
URTI: Upper Respiratory Tract Infections
IQR: Interquartile Range
CI: Confidence Interval
OR: Odds Ratio
USA: United States of America

## Acknowledgements

We thank all survivors for participating in our study, the study team of the Childhood Cancer Research Group, the data managers of the Swiss Paediatric Oncology Group, and the team of the Swiss Childhood Cancer Registry. We also thank Christopher Ritter for editorial assistance.

## Funding

This work was supported by the Swiss Cancer Research and Swiss Cancer League (Grant no. KFS-5027-02-2020, KFS-5302-02-2021, KLS/KFS-4825-01-2019, KLS/KFS-5711-01-2022), Childhood Cancer Switzerland, Kinderkrebshilfe Schweiz, Stiftung für krebskranke Kinder - Regio Basiliensis, and the University of Basel Research Fund for Excellent Junior Researchers.

## SUPPLEMENTAL LITERATURE SUMMARY

| Author, year of publication | Type of study | Country, year of study | Sample size | Age of participants | Smoking prevalence | Respiratory outcomes, OR (95% CI) | Covariates adjusted for in the analysis |
| --- | --- | --- | --- | --- | --- | --- | --- |
| **Parental smoking prevalence and associations with respiratory outcomes in healthy populations** |  |  |  |  |  |  |  |
| Gilliland, 2001 [1] | Cross-sectional, questionnaire-based | US, 1993 | 5 762 | School-aged children (no detailed information on age) | Tobacco smoke exposure<br>Active smoking by child, 1.7%<br>*In utero* smoking exposure, 18.8%<br>Any lifetime ETS exposure, 39.5%<br>Current ETS exposure, 22.4%<br>Mom only, 6.5%<br>Dad only, 6.3%<br>Both parents, 4.9%<br>Others, 4.7% | Active asthma<br>Maternal smoking, 0.9 (0.5-1.5)<br>Paternal smoking, 1.1 (0.7-1.8)<br>Wheeze with cold<br>Maternal smoking, 1.5 (1.1-2.2)<br>Paternal smoking, 1.1 (0.8-1.6)<br>Wheeze without cold<br>Maternal smoking, 1.5 (0.9-2.3)<br>Paternal smoking, 1.1 (0.7-1.8)<br>Wheeze with exercise<br>Maternal smoking, 1.7 (1.1-2.6)<br>Paternal smoking, 1.1 (0.7-1.7) | Towns, grade, cohort, age, race, sex, hay fever, family history of asthma, family history of atopy, and gestational age. |
| Strachan, 1997 and 1998 [2-4] | Systematic review, mostly questionnaire-based studies | UK, 1997 | 25 studies of asthma<br>41 of wheeze<br>34 of chronic cough<br>11 of recurrent otitis | School-aged children |  | Asthma<br>Maternal smoking, 1.36 (1.20-1.55)<br>Paternal smoking, 1.07 (0.92-1.24)<br>Wheeze<br>Maternal smoking, 1.28 (1.19-1.38)<br>Paternal smoking, 1.14 (1.06-1.23)<br>Cough<br>Maternal smoking, 1.40 (1.20-1.64)<br>Paternal smoking, 1.21 (1.09-1.34)<br>Recurrent otitis media<br>Either parent, 1.48 (1.08-2.04) |  |
| Burke, 2012 [5] | Systematic review | UK, 1997-2011 | 71 studies in total assessing asthma and/or wheeze | 5-18 years |  | Asthma<br>Maternal smoking, 1.20 (0.98-1.44)<br>Paternal smoking, 0.98 (0.71-1.36)<br>Wheeze<br>Maternal smoking, 1.18 (0.99-1.40)<br>Paternal smoking, 1.39 (1.05-1.85) |  |
| Mitchell, 2012 [6] | Cross-sectional study, questionnaire-based | multicenter, multicountry | 6-7-year age group: 220 407 children from 75 centers in 32 countries<br>13-14-year age group: 350 654 adolescents from 118 centers in 53 countries | children (aged 6-7 years) and adolescents (aged 13-14 years) |  | Current wheeze<br>Age group 6-7 years<br>Maternal smoking, 1.28 (1.22-1.34)<br>Paternal smoking, 1.17 (1.12-1.21)<br>Age group 13-14 years<br>Maternal smoking, 1.32 (1.26-1.37)<br>Paternal smoking, 1.20 (1.15-1.24)<br>Asthma ever<br>Age group 6-7 years<br>Maternal smoking, 1.26 (1.20-1.32)<br>Paternal smoking, 1.11 (1.06-1.15)<br>Age group 13-14 years<br>Maternal smoking, 1.20 (1.16-1.25)<br>Paternal smoking, 1.08 (1.04-1.12) | Gender, language, region and Gross National Income (GNI) category |
| Pattenden, 2006 [7] | Cross-sectional study, questionnaire-based | 12 countries | 53 879 | 6-12 years |  | Wheeze<br>Current parental smoking, 1.20 (1.06-1.35)<br>Asthma<br>Current parental smoking, 1.09 (1.01-1.19) | Age, sex, maternal education, paternal education, nationality, household crowding, gas for cooking, unvented gas/oil/kerosene heater, mold, birth order, "ever had a pet", and study area |
| Hur, 2013 [8] | Systematic review | US, (1998-2013) | 9/19 publications in pediatric population |  |  | Significant association between secondhand smoke and sinusitis |  |
| Jung, 2011 [9] | Cross-sectional study, questionnaire-based | Korea | 31 983 children | 6-11 years |  | Wheezing within 1 year<br>Parental secondhand smoking, 1.14 (1.06-1.22)<br>Asthma<br>Parental secondhand smoking, 1.05 (0.99-1.11)<br>Cough >3 months<br>Parental secondhand smoking, 1.21 (1.10-1.33) | Age, sex |
| Raherison, 2007 [10] | Cross-sectional study, questionnaire-based | France | 7 798 | 9-11 years | In utero<br>Mother, 21.6%<br>Father, 37.5%<br>Mother and father, 12%<br>First year of life<br>Mother, 29.8%<br>Father, 40.9%<br>Mother and father, 20%<br>Current<br>Mother, 34.1%<br>Father, 36.2%<br>Mother and father, 19.5% | Lifetime wheezing<br>Current smoking<br>Maternal, 1.02 (0.8-1.3)<br>Paternal, 0.89 (0.76-1.03)<br>Asthma<br>Current smoking<br>Maternal, 1.23 (1.02-1.47)<br>Paternal, 0.89 (0.79-1.1) | Gender, siblings, exposure to pets during the first year of life, ethnic group, parental education, day care, breast feeding, geographic center, parental atopy, prematurity |
| Tsai, 2010 [11] | Cross-sectional study, | Taiwan, 2007 | 5 804 | 12-14 years | Current smoker<br>Mother, 3.4%<br>Father, 35.7% | Asthma<br>Maternal smoking, 1.40 (0.82,2.39)<br>Paternal smoking, 1.07 (0.85,1.35) | Age, sex, parental education, family history of asthma, family history of atopy, gestational age, and community |
|  | questionnaire-based |  |  |  |  | Current wheeze<br>Maternal smoking, 2.7 (1.54,4.75)<br>Paternal smoking, 0.95 (0.69,1.32)<br>Cough >3 months<br>Maternal smoking, 2.39 (1.30,4.39)<br>Paternal smoking, 0.95 (0.68,1.33) |  |
| Vlaski, 2010 [12] | Cross-sectional study, questionnaire-based | Macedonia, 2002 | 3 024 | 13-14 years | Maternal tobacco smoke, 50.7%<br>Paternal tobacco smoke, 51.7% | Asthma<br>Maternal smoking, 1.76 (0.96-3.22)<br>Paternal smoking, 1.02 (0.57-1.83)<br>Current wheeze<br>Maternal smoking, 1.15 (0.88-1.49)<br>Paternal smoking, 1.04 (0.80-1.36) | Gender, current fruit and vegetables and cereals intake, gas/wood cooking and heating exposure at home, current cat and dog ownership, maternal educational level, siblings |
| **Respiratory outcomes in childhood cancer survivors** |  |  |  |  |  |  |  |
| Chehab, 2022 [13] | Population-based retrospective observational follow-up analysis using linked cancer-birth-hospital discharge records | USA, Washington State, 2022 | 3 152 | Children and adolescents |  | Incidence rate ratio of infections associated with a hospitalization in cancer survivors >5 years from diagnosis relative to comparators for respiratory system: 6.4 (5.1-8.0) |  |
| De Fine Licht, 2017 [14] | Retrospective, register-based cohort study | Denmark, Finland, Iceland, and Sweden | 21,297 5-year survivors, 152,231 population comparison | Any age >5 years |  | Standardized hospitalization rate ratios (RRs):<br>Respiratory system diseases, 1.6 (1.5–1.7)<br>- Acute upper respiratory infections 1.3 (1.2–1.4) |  |
| Dietz, 2016 [15] | Cross-sectional, questionnaire-based study | USA | 20 690 | Median age at study: 32 years (range, 6-59 years) |  | Asthma 7.2%<br>Chronic cough 7.8%<br>Rate ratio for survivors vs siblings:<br>- Asthma 1.0 (0.9-1.2)<br>- Chronic cough 1.6 (1.4-1.9) |  |
| Mertens, 2002 [16] | Cross-sectional, questionnaire-based study | USA | 12 390 survivors and 3546 siblings | Any age (participants <18y: 33.2%) |  | Rate ratio compared to siblings:<br>- Chronic cough, 2.3 (1.9-2.8)<br>- Recurrent sinusitis, 1.7 (1.5-1.9)<br>- Asthma, 1.3 (1.1-1.5) |  |
| Mueller, 2018 [17] | Case-control study | USA | 3152 survivors, 31520 comparison children | 5-39 years |  | HR hazard ratio for any hospitalization, death (any cause), and cause-specific hospitalization/death (combined):<br>Respiratory system, 4.0 (3.3-4.9)<br>Infections, 3.2 (2.7-3.8) |  |
| Perkins, 2014 [18] | Cross-sectional, questionnaire-based study | USA, 1994-2007 | 12 360 | Any age (participants <20y: 8.6%) |  | Sinusitis, 13.5%<br>Chronic cough, 7.2%<br>Rate ratio relative to siblings:<br>- Sinusitis, 1.8 (1.6-1.9) | Patients who reported being current tobacco smokers (RR, 1.4; 95% CI, 1.2-1.5) or former tobacco smokers |
|  |  |  |  |  |  | - Chronic cough, 2.0 (1.8-2.3) | (RR, 1.3; 95% CI, 1.1-1.5) had elevated rates of sinopulmonary infection compared with those who reported having never smoked. |
| **Passive smoking exposure in childhood cancer survivors** |  |  |  |  |  |  |  |
| Chung, 2022 [19] | Cross-sectional, questionnaire-based study | Hong Kong | 200 (pediatric survivors n=45) | The mean (standard deviation) ages at interview were 26.9 (6.4) years and 11.1 (3.6) years among adult and pediatric survivors |  | Exposure to second-hand smoking (adult and pediatric survivors):<br>Yes 85 (42.5%)<br>- Family (residing in the same household) 41 (20.5%)<br>- Friends 27 (13.5%)<br>- Colleagues 29 (14.5%)<br>- Others 11 (5.5%) |  |
| Cho, 2020 [20] | Case-control study, questionnaire-based | South Korea | 326 caregivers of childhood cancer survivors (parents of 240 CCS) |  |  | Current smoking:<br>All subjects, 15%<br>Male (fathers), 34.7%<br>Female (mothers), 3.4% |  |
| Badr, 2011 [21] | Cross-sectional, questionnaire-based study | USA, Texas, 2009 | 170 survivors and 114 parents | Mean age of survivors at study: 17.8 years (SD 8.3 years)<br>Average age of parents: 42.6 years (SD 7.7 years) |  | Survivors:<br>- current smokers 7.4%, n=10<br>- recent quitters 5.2%, n=7<br>Parents:<br>- current smokers (6.3%, n=7)<br>- recent quitters (20.5%, n=23) |  |

**SUPPLEMENTAL FIGURE S1.**
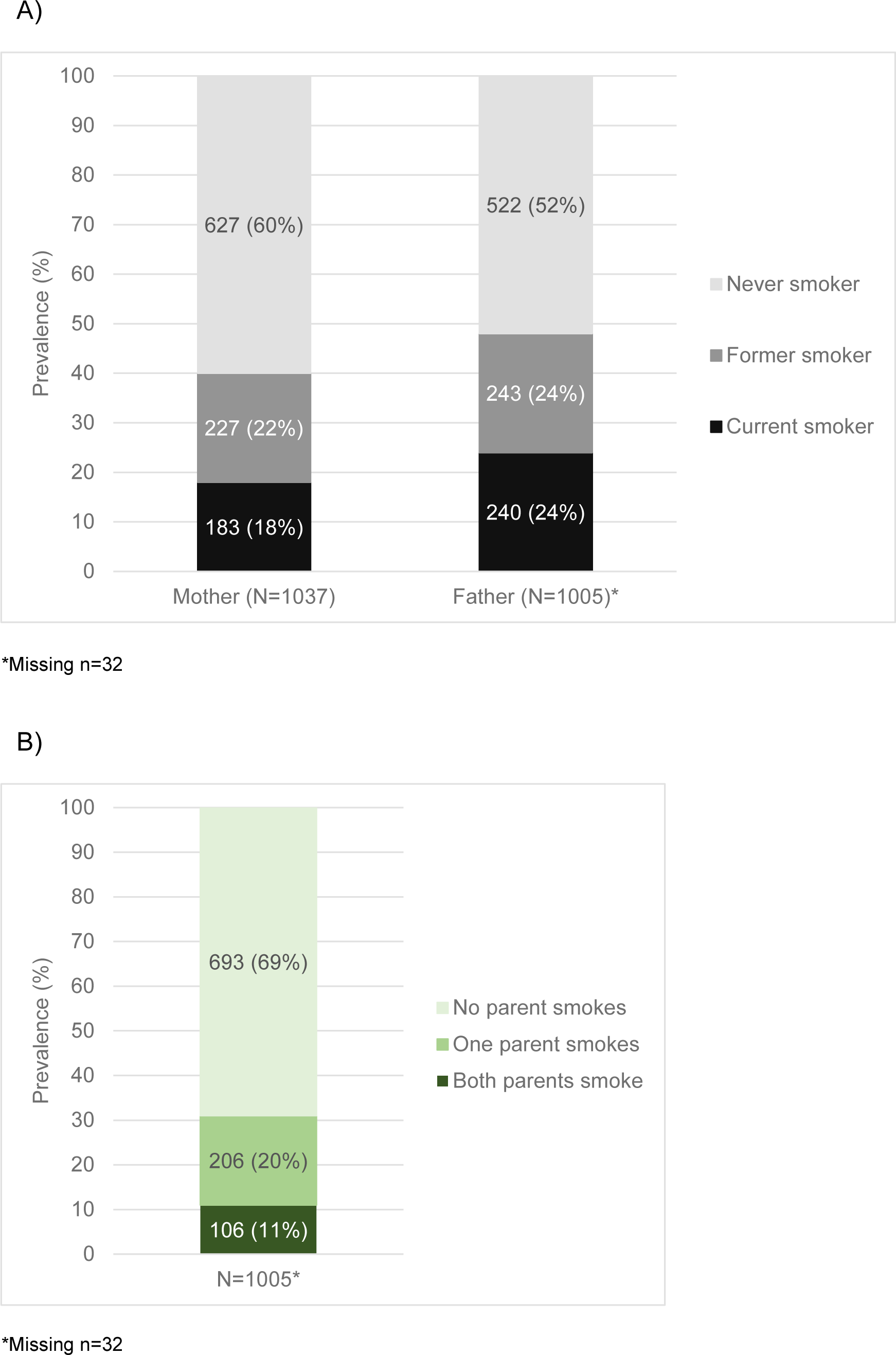
Proportion of mothers and fathers of childhood cancer survivors who were active smokers (A) and proportion of families with one or two parents who were active smokers.

**SUPPLEMENTAL TABLE S1.** Characteristics of mothers and fathers of childhood cancer survivors from the Swiss Childhood Cancer Survivor Study.

| N=1037 |  |  |  |  |
| --- | --- | --- | --- | --- |
| Sociodemographic characteristics | Mothers |  | Fathers |  |
|  | n | (%) | n | (%) |
| *Median age at survey in years (IQR)* | 43 (40–47) |  | 46 (42–50) |  |
| *Nationality* |  |  |  |  |
| Swiss | 742 | (72) | 740 | (71) |
| Other | 283 | (27) | 266 | (26) |
| Unknown (missing) | 12 | (1) | 31 | (3) |
| *Education (highest level)* |  |  |  |  |
| Primary education | 132 | (13) | 93 | (9) |
| Secondary education | 515 | (50) | 372 | (36) |
| Tertiary education | 363 | (35) | 506 | (49) |
| Unknown (missing) | 27 | (2) | 66 | (6) |
| *Employment status* |  |  |  |  |
| Full-time job | 106 | (10) | 832 | (80) |
| Part-time job | 532 | (51) | 63 | (6) |
| Several part-time jobs | 101 | (10) | 22 | (2) |
| Unemployed<sup>a</sup> | 241 | (23) | 31 | (3) |
| Unknown (missing) | 57 | (6) | 89 | (9) |
| *Smoking status* | n | (%) | n | (%) |
| Never smoker | 627 | (60) | 522 | (50) |
| Former smoker | 227 | (22) | 243 | (24) |
| Current smoker | 183 | (18) | 240 | (23) |
| Unknown (missing) | 0 | (0) | 32 | (3) |

<sup>a</sup> Including parents who are in education.
Abbreviations: IQR, inter quartile range.

**SUPPLEMENTAL TABLE S2.**
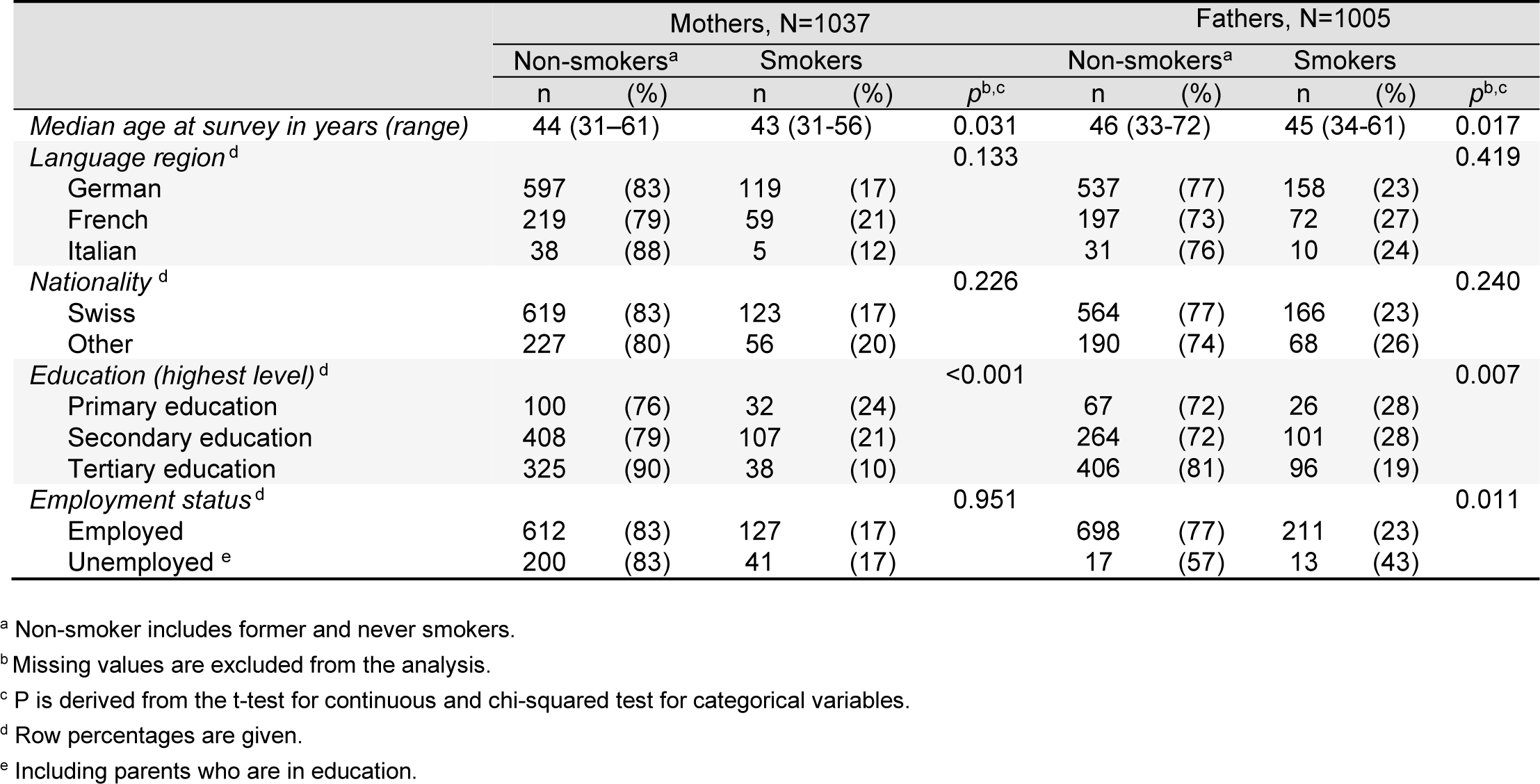
Comparison of characteristics of non-smokers and smokers among mothers and fathers of childhood cancer survivors.

|  | Mothers, N=1037 |  |  |  |  | Fathers, N=1005 |  |  |  |  |
| --- | --- | --- | --- | --- | --- | --- | --- | --- | --- | --- |
|  | Non-smokers<sup>a</sup> |  | Smokers |  | *p*<sup>b,c</sup> | Non-smokers<sup>a</sup> |  | Smokers |  | *p*<sup>b,c</sup> |
|  | n | (%) | n | (%) |  | n | (%) | n | (%) |  |
| *Median age at survey in years (range)* | 44 (31–61) |  | 43 (31–56) |  | 0.031 | 46 (33–72) |  | 45 (34–61) |  | 0.017 |
| *Language region<sup>d</sup>* |  |  |  |  | 0.133 |  |  |  |  | 0.419 |
| German | 597 | (83) | 119 | (17) |  | 537 | (77) | 158 | (23) |  |
| French | 219 | (79) | 59 | (21) |  | 197 | (73) | 72 | (27) |  |
| Italian | 38 | (88) | 5 | (12) |  | 31 | (76) | 10 | (24) |  |
| *Nationality<sup>d</sup>* |  |  |  |  | 0.226 |  |  |  |  | 0.240 |
| Swiss | 619 | (83) | 123 | (17) |  | 564 | (77) | 166 | (23) |  |
| Other | 227 | (80) | 56 | (20) |  | 190 | (74) | 68 | (26) |  |
| *Education (highest level)<sup>d</sup>* |  |  |  |  | <0.001 |  |  |  |  | 0.007 |
| Primary education | 100 | (76) | 32 | (24) |  | 67 | (72) | 26 | (28) |  |
| Secondary education | 408 | (79) | 107 | (21) |  | 264 | (72) | 101 | (28) |  |
| Tertiary education | 325 | (90) | 38 | (10) |  | 406 | (81) | 96 | (19) |  |
| *Employment status<sup>d</sup>* |  |  |  |  | 0.951 |  |  |  |  | 0.011 |
| Employed | 612 | (83) | 127 | (17) |  | 698 | (77) | 211 | (23) |  |
| Unemployed<sup>e</sup> | 200 | (83) | 41 | (17) |  | 17 | (57) | 13 | (43) |  |

<sup>a</sup> Non-smoker includes former and never smokers.
<sup>b</sup> Missing values are excluded from the analysis.
<sup>c</sup> P is derived from the t-test for continuous and chi-squared test for categorical variables.
<sup>d</sup> Row percentages are given.
<sup>e</sup> Including parents who are in education.

**SUPPLEMENTAL TABLE S3.** Multivariable logistic regression analysis between parental smoking and respiratory outcomes in childhood cancer survivors. Models are adjusted for age at survey, sex, and shown treatment-related factors.

|  | Recurrent URTI<sup>a</sup> |  |  | Asthma<sup>a</sup> |  |  | Lower respiratory symptoms<sup>a</sup> |  |  |
| --- | --- | --- | --- | --- | --- | --- | --- | --- | --- |
|  | OR | (95% CI) | p<sup>b</sup> | OR | (95% CI) | p<sup>b</sup> | OR | (95% CI) | p<sup>b</sup> |
| *Mother's smoking status* |  |  | 0.016 |  |  | 0.444 |  |  | 0.035 |
| Non-smoker<sup>c</sup> | Ref. |  |  | Ref. |  |  | Ref. |  |  |
| Smoker | 2.1 | (1.1-3.7) |  | 1.2 | (0.6-2.6) |  | 2.0 | (1.1-3.7) |  |
| *Father's smoking status* |  |  | 0.199 |  |  | 0.822 |  |  | 0.692 |
| Non-smoker<sup>c</sup> | Ref. |  |  | Ref. |  |  | Ref. |  |  |
| Smoker | 0.7 | (0.4-1.2) |  | 1.1 | (0.5-2.2) |  | 1.1 | (0.6-2.1) |  |
| *Age at survey* | 0.9 | (0.8-1.0) | 0.037 | 1.0 | (0.9-1.1) | 0.983 | 1.0 | (0.9-1.1) | 0.999 |
| *Sex* |  |  | 0.733 |  |  | 0.108 |  |  | 0.534 |
| Male | Ref. |  |  | Ref. |  |  | Ref. |  |  |
| Female | 1.1 | (0.7-1.7) |  | 0.6 | (0.3-1.1) |  | 0.8 | (0.5-1.4) |  |
| *Time since diagnosis* | 0.9 | (0.8-1.1) | 0.410 | ‡ |  |  | 0.9 | (0.8-1.0) | 0.082 |
| *Relapse* | ‡ |  |  | ‡ |  |  |  |  | 0.052 |
| No |  |  |  |  |  |  | Ref. |  |  |
| Yes |  |  |  |  |  |  | 1.9 | (1.0-3.8) |  |

The respective multivariable models were adjusted for variables with a p-value of 0.1 or less in the univariable logistic regression.
<sup>a</sup> Participants with missing data for paternal smoking were excluded from the logistic regression models.
<sup>b</sup> P calculated from Wald test.
<sup>c</sup> Non-smoker includes former and never smokers.
‡ Not included in multivariable logistic regression as p>0.1 in univariable logistic regression
Abbreviations: URTI, upper respiratory tract infections; OR, odds ratio; CI, confidence interval.

**SUPPLEMENTAL TABLE S4.** Univariable logistic regression analysis between parental smoking, sociodemographic characteristics, and cancer-related characteristics, and respiratory health outcomes in childhood cancer survivors.

|  | Recurrent URTI |  |  | Asthma |  |  | Lower respiratory symptoms |  |  |
| --- | --- | --- | --- | --- | --- | --- | --- | --- | --- |
|  | OR | (95% CI) | *p*<sup>a</sup> | OR | (95% CI) | *p*<sup>a</sup> | OR | (95% CI) | *p*<sup>a</sup> |
| **Sex** |  |  | 0.754 |  |  | 0.055 |  |  | 0.475 |
| Male | Ref. |  |  | Ref. |  |  | Ref. |  |  |
| Female | 1.1 | (0.7-1.7) |  | 0.6 | (0.3-1.0) |  | 0.8 | (0.5-1.4) |  |
| **Age at survey (years)** | 0.9 | (0.8-1.0) | 0.008 | 1.0 | (0.9-1.1) | 0.904 | 1.0 | (0.9-1.1) | 0.329 |
| **Mother's smoking status** |  |  | 0.040 |  |  | 0.367 |  |  | 0.005 |
| Non-smoker<sup>b</sup> | Ref. |  |  | Ref. |  |  | Ref. |  |  |
| Smoker | 1.7 | (1.0-2.9) |  | 1.4 | (0.7-2.6) |  | 2.2 | (1.3-3.8) |  |
| **Father's smoking status** |  |  | 0.875 |  |  | 0.598 |  |  | 0.155 |
| Non-smoker<sup>b</sup> | Ref. |  |  | Ref. |  |  | Ref. |  |  |
| Smoker | 1.0 | (0.6-1.6) |  | 1.2 | (0.6-2.2) |  | 1.5 | (0.9-2.6) |  |
| **Migration background** |  |  | 0.133 |  |  | 0.916 |  |  | 0.491 |
| No | Ref. |  |  | Ref. |  |  | Ref. |  |  |
| Yes | 0.7 | (0.4-1.1) |  | 1.0 | (0.6-1.8) |  | 1.2 | (0.7-2.0) |  |
| **Swiss language region** |  |  | 0.165 |  |  | 0.931 |  |  | 0.756 |
| German | Ref. |  |  | Ref. |  |  | Ref. |  |  |
| French or Italian | 1.4 | (0.9-2.2) |  | 1.0 | (0.6-1.9) |  | 0.9 | (0.5-1.6) |  |
| **Parent's education (highest level)<sup>c</sup>** |  |  | 0.429 |  |  | 0.587 |  |  | 0.839 |
| Primary education | 0.5 | (0.2-1.5) |  | 1.6 | (0.6-4.1) |  | 0.8 | (0.3-2.2) |  |
| Secondary education | Ref. | (0.7-3.2) |  | Ref. |  |  | Ref. |  |  |
| Tertiary education | 0.8 | (0.5-1.3) |  | 1.0 | (0.5-1.8) |  | 0.9 | (0.5-1.5) |  |
| **Time since diagnosis** | 0.9 | (0.8-1.0) | 0.027 | 1.0 | (0.9-1.1) | 0.923 | 0.9 | (0.8-1.0) | 0.065 |
| **Chemotherapy** |  |  | 0.399 |  |  | 0.563 |  |  | 0.650 |
| No | Ref. |  |  | Ref. |  |  | Ref. |  |  |
| Yes | 1.3 | (0.7-2.5) |  | 0.8 | (0.4-1.6) |  | 1.2 | (0.6-2.3) |  |
| **Radiation** |  |  | 0.291 |  |  | 0.207 |  |  | 0.565 |
| No | Ref. |  |  | Ref. |  |  | Ref. |  |  |
| Yes | 0.7 | (0.4-1.3) |  | 0.6 | (0.3-1.3) |  | 0.8 | (0.4-1.6) |  |
| **Surgery** |  |  | 0.416 |  |  | 0.112 |  |  | 0.120 |
| No | Ref. |  |  | Ref. |  |  | Ref. |  |  |
| Yes | 0.8 | (0.5-1.3) |  | 0.2 | (0.1-1.5) |  | 0.7 | (0.4-1.1) |  |
| **HSCT** |  |  | 0.697 |  |  | 0.710 |  |  | 0.856 |
| No | Ref. |  |  | Ref. |  |  | Ref. |  |  |
| Yes | 1.2 | (0.5-2.5) |  | 0.9 | (0.5-1.6) |  | 0.9 | (0.4-2.3) |  |
| **Relapse** |  |  | 0.126 |  |  | 0.707 |  |  | 0.027 |
| No | Ref. |  |  | Ref. |  |  | Ref. |  |  |
| Yes | 1.6 | (0.9-3.0) |  | 1.2 | (0.5-2.7) |  | 2.1 | (1.1-3.9) |  |

<sup>a</sup> P calculated from Wald test.
<sup>b</sup> Non-smoker includes former and never smokers.
<sup>c</sup> The highest completed education of either father or mother.
Abbreviations: URTI, upper respiratory tract infections; OR, odds ratio; CI, confidence interval; HSCT, hematopoietic stem cell transplantation

## Notes

### Competing Interest Statement

The authors have declared no competing interest.

### Funding Statement

This study was supported by the Swiss Cancer Research and Swiss Cancer League (Grant no. KFS-5027-02-2020, KFS-5302-02-2021, KLS/KFS-4825-01-2019, KLS/KFS- 5711-01-2022), Childhood Cancer Switzerland, Kinderkrebshilfe Schweiz, Stiftung fuer krebskranke Kinder - Regio Basiliensis, and the University of Basel Research Fund for Excellent Junior Researchers.

### Author Declarations

The Ethics Committee of the Canton of Bern granted ethical approval to the Swiss Childhood Cancer Registry and the Swiss Childhood Cancer Survivor Study (166/2014; 2021-0146).

